# Effect of Ozone on Allergic Airway Inflammation

**DOI:** 10.1101/2022.01.05.22268824

**Authors:** Mehrdad Arjomandi, Hofer Wong, Rachel Tenney, Nina Holland, John R. Balmes

**Affiliations:** Department of Medicine, University of California, San Francisco; Medical Service San Francisco Veterans Affairs Healthcare System; Division of Environmental Health Sciences, School of Public Health, University of California, Berkeley

**Keywords:** Ozone, allergen, airway inflammation, local endobronchial allergen challenge, neutrophils, cytokines, glutathione s-transferase mu (GSTM1)

## Abstract

**Background:** Exposure to O_3_ has been associated with increased risk of exacerbations of asthma, but the underlying mechanisms are not well studied. We hypothesized that O_3_ exposure would enhance airway inflammatory responses to allergen and the GSTM1 null genotype would modulate this enhancement.

**Procedures:** In a cross-over design, 10 asthmatic subjects (50% with GSTM1 null genotype) who had specific sensitization to *Dermatophagoides pteronyssinus* (DP) were exposed to 160 ppb O_3_ or filtered air (FA) control for 4 h with intermittent exercise on two separate days at least three weeks apart. 20 h post-exposure, endobronchial challenge with DP allergen, and sham normal saline (NS) instillation, were performed in two separate lung lobes. Six h later, a second bronchoscopy was performed to collect bronchoalveolar lavage (BAL) fluid from the DP- and NS-challenged lobes for analyses of cellular and biochemical markers of inflammation. Multiple variable regression was used to compare cell and cytokine responses across the four exposure groups (FA-NS, O_3_-NS, FA-DP, O_3_-DP). Effect modification by GSTM1 genotype was assessed in stratified regressions.

**Main Findings:** BAL eosinophil and lymphocyte counts were increased in segments challenged with DP compared to segments that received sham challenges (p<0.01). DP challenge compared to sham challenge also caused a significant increase in BAL concentrations of the Th2 cytokines IL-4, IL-5, IL-10, and IL-13 (p<0.03 for all comparisons). O_3_ exposure did not significantly affect BAL cells or cytokine levels although BAL neutrophil count with DP challenge was non-significantly higher after O_3_ compared to after FA exposure (p<0.11). Compared to GSTM1-present subjects, GSTM1-null subjects had significantly reduced inflammatory responses including lower eosinophil (p<0.041) and IL-4 (p<0.014) responses to DP challenge after O_3_ exposure.

**Conclusions:** O_3_ appears to have mixed effects on allergen-induced airway inflammation. While O_3_ did not cause a clear differential effect on airway cellular or cytokine responses to allergen challenge, those responses did appear to be modulated by the antioxidant enzyme, GSTM1, as evident by the attenuation of airway inflammatory responses to allergen after O_3_ exposure in the absence of the gene.

**HIGHLIGHTS:**

- Ozone may increase risk of asthma exacerbation but the exact mechanisms are not clear.
- Susceptibility to ozone-induced airway inflammation may be associated with GSTM1 genotype.
- Ozone may enhance allergen-induced airway recruitment of neutrophils.
- The GSTM1 null mutation may decrease both eosinophil and cytokine allergic airway responses after O_3_ exposure.

## INTRODUCTION

Ozone (O_3_) is a major gaseous component of air pollution in many countries. Epidemiological evidence suggests that people with asthma are at increased risk for exacerbation when exposed to elevated levels of ambient O_3_ (1). Controlled human exposure studies have not consistently shown subjects with asthma to be more sensitive to O_3_ in terms of lung function response, although the neutrophilic airway inflammatory response does appear to be greater in asthmatic than in non-asthmatic subjects (2, 3). In addition, there is evidence that lung function and airway inflammatory responses to O_3_ are not well-correlated in healthy subjects (4). Asthma is a disease characterized by airway inflammation, particularly during the late-phase response to allergen, and the degree of airway inflammation is an important predictor of asthma severity. Thus, one possible explanation for the epidemiological findings is that O_3_ exposure may enhance the inflammatory response to triggers of asthma, such as allergen, not reflected in prior controlled human studies measuring lung function 1parameters alone.

Animal toxicological data provide evidence that O_3_ exposure can enhance allergic inflammatory responses in the lungs (5, 6), but at least one study in a dog model showed that O_3_ pre-exposure attenuated the late-phase response to sub-lobar placement of antigen (7). Controlled human exposure studies have confirmed that O_3_ exposure can enhance both the early and late bronchoconstrictor responses to inhaled antigen in some, but not all allergic asthmatic subjects (8, 9). Unlike bronchoconstriction, the effect of O_3_ on allergen-induced airway inflammation has not been well studied, and most of the published studies did not assess potential changes in airway inflammation during the late-phase response. However, in the two studies that did, significant O_3_- induced enhancement was not consistently observed (10, 11).

Ozone is a prototypic oxidant pollutant that can generate reactive oxygen species (ROS) in the airways when inhaled, potentially leading to oxidative stress. Although innate antioxidant defenses are available to detoxify ROS in the airway, individuals differ in their ability to deal with an oxidant burden, such as inhaled O_3_, and such differences are in part genetically determined. Decreased ability to detoxify ROS may lead to enhanced airway inflammation, and thus potentially to increased bronchoconstriction and asthma symptoms. The glutathione S-transferase (GST) enzymes comprise a large supergene family located on at least seven chromosomes that are critical to the protection of cells from ROS (12). GSTM1 is a polymorphic gene with a common null allele (13, 14). The null allele is unable to produce a functional enzyme, which would in turn be expected to affect response to oxidative stress. From 30-65% of the general population is GSTM1 null (15). The results of several controlled human exposure studies have suggested that individuals who are GSTM1 null have greater lung function responses to O_3_ exposure compared to individuals with the form of the gene that produces functional enzyme (16, 17). However, two studies did not demonstrate an effect of GSTM1 status on the airway inflammatory and lung function responses to O_3_ in both asthmatic and non-asthmatic adult subjects (18, 19). A third study of non-asthmatic subjects using a higher O_3_ concentration (400 ppb) did show that the GSTM1 null genotype was associated with increased airways inflammation 24 hours after exposure (20).

Based on the previous work indicating that O_3_ enhanced the physiologic responses to inhaled allergen, we hypothesized that O_3_ exposure would also enhance allergic airway inflammation. To test this hypothesis, we conducted a controlled human exposure study with a repeated measure crossover design that used O_3_ or FA exposure prior to administration of local endobronchial allergen challenge (LEAC) with DP and saline in different lobes of the lungs. We also hypothesized that the effects of inhaled O_3_ on the specific airway inflammatory responses to allergen would be enhanced in asthmatic individuals with the GSTM1 null genotype compared to those who have the functional form of the GSTM1 gene.

## METHODS

### Study Design

This study had a repeated measure design in which specifically-sensitized asthmatic participants were exposed to either clean filtered air (FA) or 160 ppb for 4 hours in a climate-controlled chamber followed by a challenge bronchoscopy approximately 20 hours later and a sampling bronchoscopy 6 hours after the endobronchial challenge. Spirometry was performed immediately before exposure (0-h), immediately after exposure (4-h), and on the following morning prior to bronchoscopy (24-h). In addition, spirometry was performed on an hourly basis after the challenge bronchoscopy through discharge of the participant approximately 2 h after the sampling bronchoscopy. Each participant returned and underwent the second exposure type with a minimum of 2 weeks in between exposure sessions to allow for recovery from any inflammation or injury sustained during the prior session. The order of exposures was counterbalanced and randomized. The investigators did not know the GSTM1 genotype of participants during data collection.

### Participants

The inclusion/exclusion criteria included: (1) age between 18 to 50 years; (2) ability to perform moderate-intensity exercise; (3) being healthy with no history of cardiovascular, hematologic, or pulmonary diseases other than mild asthma; (4) specific sensitization to the house dust mite, *Dermatophagoides pteronyssinus* (DP); (5) no history of acute infection within the 6 weeks prior to start of the study; (6) non-smoker as defined as having a history of less than ½ pack-year lifetime tobacco use and no history of any tobacco use in the past 6 months; and (7) no history of illicit drug use. The participants were asked to stop their asthma and allergy medications in a sequential manner based on the duration of action of each medication (inhaled corticosteroids for 2 weeks, anti-histamines and leukotriene inhibitors for 3 days, long-acting bronchodilators for 2 days, and short-acting bronchodilators for 8 h). The participants were informed of the risks of the experimental protocol and signed a consent form that had been approved by the University of California San Francisco (UCSF) Committee on Human Research. All participants received financial compensation for their participation.

Ten partcipants were recruited via advertisements placed in campus newsletters, local San Francisco newspapers, and internet websites (e.g., www.craigslist.org). A total of 542 individuals responded to the Craig’s List postings and all were contacted by email. Of these, 20 consented to participate in the study, of which 10 completed the study. Of the other 10, one was discontinued because of a severe hypotensive episode with syncope secondary to anaphylaxis, one was ineligible due to lack of airway hyperresponsiveness, three were lost to follow-up, two withdrew consent due to work scheduling issues, one was ineligible due to a pulmonary interstitial lung disease diagnosis, and two were ineligible due to a negative DP skin test.

### Allergy Skin Testing

(Pre-enrollment) To determine allergy status, and sensitivity to *Dermatophagoides pteronyssinus* (DP) an allergy skin testing with a set of 10 common aeroallergens [DP, birch mix, chinese elm, cat, dog, mountain cedar, mugwort sage, olive tree, perennial rye, *aspergillus fumigatus*] and controls of saline and histamine was performed inside the forearm. Sensitivity was defined as a >2 x 2 mm skin wheal response, except for DP (>3 x 3 mm skin wheal). If the subject was sensitive to DP on the initial skin-prick test, a dilutional skin test using log concentrations (1.5 AU to 15,000 AU) of DP allergen was also be performed, to determine the dose of DP allergen to be used for the allergen bronchoscopy.

### Methacholine Challenge Testing

(Pre-enrollment) To assess asthma status, a methacholine inhalation test was be performed following a protocol modified from the American Thoracic Society guidelines (21), using a nebulizer (DeVilbiss) and dosimeter (Rosenthal) set to deliver 9 µL per breath. Participants inhaled aerosol from the nebulizer in five breaths, (one every 12 seconds over a 1-minute period) and spirometry was measured 3 min after each dose. The next dose was administered within 30 seconds of completing the spirometry. Increasing doses of methacholine (0.0625, 0.125, 0.25, 0.5, 1, 2, 4, 8 mg/mL) were given, until a 20% decrease in FEV_1_ from saline FEV_1_ was achieved. A positive methacholine test was defined as a 20% decrease in FEV_1_ at <8 mg/mL.

### Climate-Controlled Chamber and Atmospheric Monitoring

The experiments took place in a ventilated, climate-controlled chamber at 20°C and 50% relative humidity. The chamber is a stainless steel-and-glass room of 2.5 × 2.5 × 2.4 m (Model W00327-3R; Nor-Lake, Hudson, WI) that was custom-built and designed to maintain temperature and relative humidity within 2.0°C and 4% from the set points, respectively (WebCtrl Software; Automated Logic Corporation, Kennesaw, GA). Temperature and relative humidity were recorded every 30 s and displayed in real-time (LabView 6.1; National Instruments, Austin, TX).

### Exposure Session

After a telephone interview, partcipants were scheduled for an initial visit to the laboratory, where a medical history questionnaire was completed. A 30-min exercise test designed to determine a workload that generated the target ventilatory rate was also completed on the initial visit. Each exposure session was 4 h long, with partcipants exercising for the first 30 min and then resting for the following 30 min of each hour in the climate-controlled chamber. The exercise consisted of running on a treadmill or pedaling a cycle ergometer. Exercise intensity was adjusted for each subject to achieve a target expired minute ventilation of 20 L/min/m^2^ body surface area. During exercise, VE was calculated (LabView 6.1; National Instruments, Austin, TX) from tidal volume and breathing frequency measured using a pneumotachograph at the 10-min and 20-min intervals of each 30-min exercise period. Participants remained inside the chamber for the entire 4-h exposure period. The type of exposure (FA or O_3_) was chosen randomly prior to each session and was not revealed to the participants.

### Spirometry

Each participant’s spirometry and peak expiratory flow were measured at each of the 0-h, 4-h, and 24-h time points. Spirometry was performed on a dry rolling-seal spirometer (S&M Instruments, Louisville, CA) following American Thoracic Society (ATS) performance criteria (22). The best values for FVC and FEV_1_ from three acceptable FVC maneuvers were used in data analysis. After the challenge bronchoscopy, the participants performed spirometry on an hourly basis using a portable spirometer (EasyOne, ndd Medical Technologies Inc., Andover, MA), again according to ATS performance criteria.

### Bronchoscopy, Endobronchial Allergen Challenge, and Lavage Procedures

The technique of local endobronchial allergen challenge (LEAC) has been shown to be safer and more effective at inducing a measurable allergic airway inflammatory response than whole lung inhalational challenge because bronchoconstriction is localized and a relatively larger amount of allergen can be delivered to the challenged lung segment and a second lung segment can be sham-challenged with saline (23, 24).

DP allergen for LEAC was obtained from Hollister-Stier Laboratories, LLC, Spokane, WA. An Investigational New Drug (IND) application for non-approved use of DP allergen manufactured for skin prick testing was filed and approved with the Food and Drug Administration (BB-IND 13354).

Allergen challenge bronchoscopies were performed 20 ± 2 h after exposure. This time was chosen because previous studies have documented the presence of an ozone-induced inflammatory response in many partcipants at this time point (25). Our laboratory’s procedures of bronchoscopy and bronchoalveolar lavage (BAL) have been previously discussed in detail (25). Briefly, intravenous access was established, supplemental O_2_ was delivered, and the upper airways were anesthetized with topical lidocaine. Sedation with intravenous midazolam and fentanyl was used as needed for participant comfort. In addition, the LEAC bronchoscopies were conducted according to the guidelines of the European Respiratory Society (24). The bronchoscope was first directed into the right upper lobe anterior segment orifice (RUL), where a control challenge was performed with 20 mL of sterile 0.9% saline (normal saline, NS) pre-warmed to 37°C. The bronchoscope was then advanced to the right middle lobe medial segment orifice (RML), where the allergen challenge was performed with 20 mL of pre-warmed DP allergen solution. The concentration of DP chosen for LEAC was 1/10 the dilution that elicited a 3-mm diameter skin wheal response. The exact concentrations of DP allergen used are shown in Supplemental Table 1. The bronchoscope was then withdrawn and the participant taken back to the clinical research center for monitoring and recovery. After the challenge bronchoscopy, the participant was monitored continuously and underwent hourly spirometry prior to the sampling bronchoscopy.

The sampling bronchoscopy was performed 6 h after the challenge bronchoscopy. The bronchoscope was first directed into the RUL where lavage was performed with two 50-ml aliquots of NS warmed to 37° C. The bronchoscope was then directed to the RUL where again lavage was performed with two 50-ml aliquots of NS warmed to 37° C. The RUL and RML fluids returned were immediately put on ice. After the sampling bronchoscopy, the participant was observed for an approximate 2-h recovery period.

Total cells were counted on uncentrifuged aliquots of BAL using a hemocytometer. Differential cell counts were obtained from slides prepared using a cytocentrifuge, 25 g for 5 min, and stained with Diff-Quik as previously described (25). Cells were counted by two independent observers; the average of the two counts was used in data analysis. BAL fluid was then centrifuged at 180 g for 15 min, and the supernatant was separated and re-centrifuged at 1,200 g for 15 min to remove any cellular debris prior to freezing at -80° C.

Concentrations of BAL cytokines were measured using a Milliplex human 9-plex cytokine assay (Millipore Corporation, St. Charles, MO). Cytokines measured included the following: granulocyte macrophage colony-stimulating factor (GM-CSF), interleukin-1β (IL-1β), interleukin 4 (IL-4), interleukin 5 (IL-5), interleukin 6 (IL-6), interleukin 7 (IL-7), interleukin 8 (IL-8), interleukin 10 (IL-10), interleukin 13 (IL-13), and tumor necrosis factor alpha (TNF-α). The lower limit of detection for GM-CSF, IL-1β, IL-4, IL-5, IL-6, IL-10, IL-13, and TNF-α was 3.2 pg/ml and for IL-8 was 16.0 pg/ml.

### GSTM1 Genotyping

DNA was isolated from whole blood using a Qiamp Blood DNA Maxi kit (Qiagen, Inc., Santa Clarita, CA). The assessment of GSTM1 genotype was done by multiplex polymerase chain reaction (PCR) using the following primers: 5’-CTGGATTGTAGCAGATCATGC-3’ and 5’-TACTTGATTGATGGGGCTCAC-3’. Briefly, 100 ng of DNA was added to 50 uL reaction containing 0.1 uM of primers, 0.2 mM each dNTP, 2.5 units of Taq polymerase, and 1.5mM magnesium chloride. Amplification was performed up to 40 steps. Products for the polymorphisms were identified on 3.5% agarose gel.

### Data Management and Statistical Analysis

All data were entered into a database (Microsoft Excel 2003; Microsoft; Redmond, WA) and then analyzed using STATA statistics software (STATA IE, version 14.0; StataCorp; College Station, TX). Student’s t-test was used for initial pair-wise comparisons of spirometric and between the two exposure types. The change in spirometric parameters over the course of each exposure was calculated linearly using the 0-h value as the baseline. Each subject served as their own control. Data are presented as mean±SD. Multiple variable regression was used to compare cell and cytokine responses across the four exposure groups (FA-NS, O_3_-NS, FA-DP, O_3_-DP). Effect modification by GSTM1 genotype was assessed in stratified regressions. A p-value of <0.05 was considered to be statistically significant in all analyses.

## RESULTS

### Participant Characteristics

Participant characteristics are shown in Table 1. Of the 10 participants who completed the study protocol, all 10 had mild asthma. Five were GSTM1-present and five were GSTM1-null. The two GSTM1-genotype groups were similar except the GSTM1-null group was older and had a higher mean BMI.

**Table 1:**
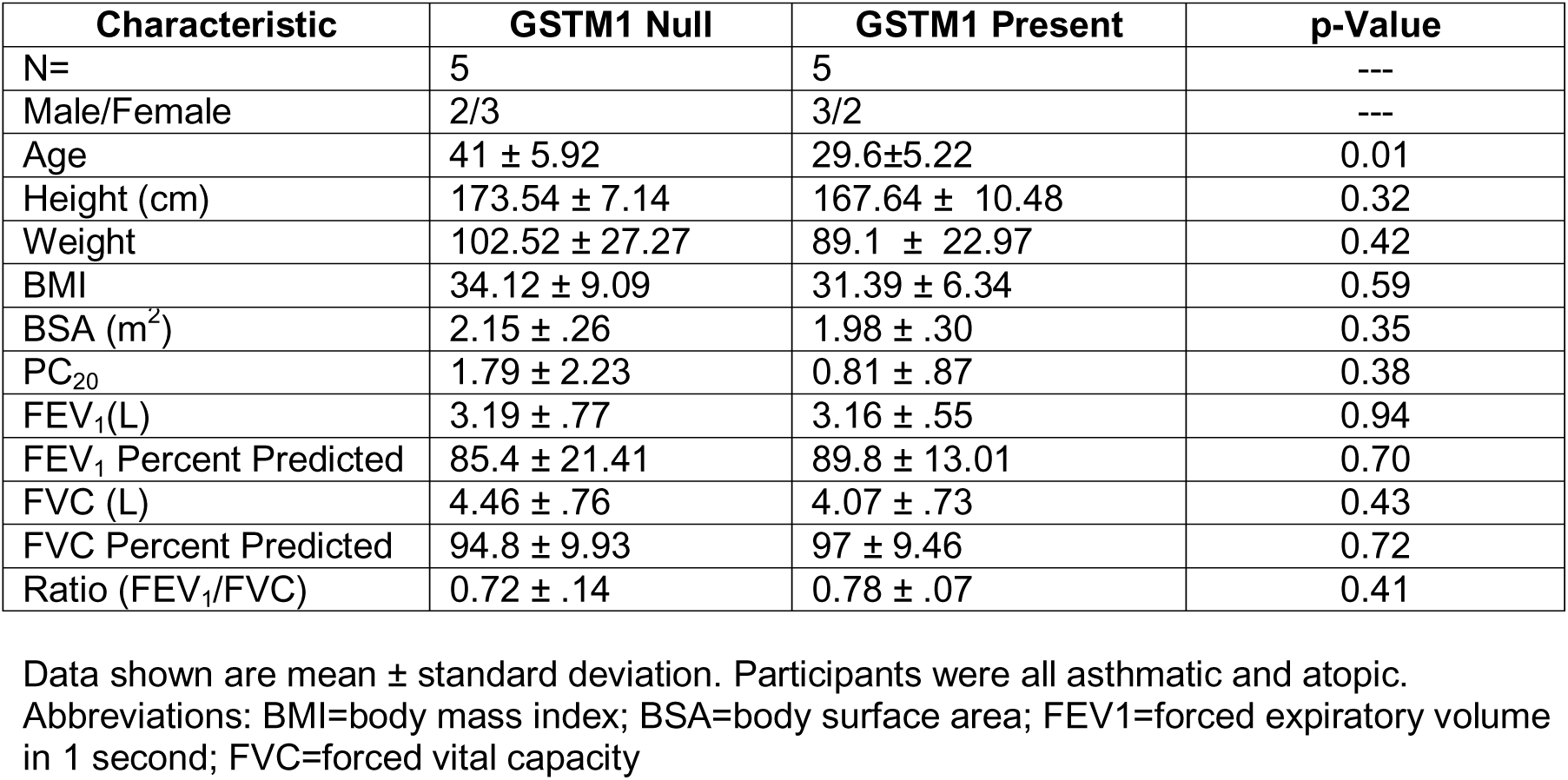
Baseline characteristics of participants

### Climate-Controlled Chamber Conditions

The temperature and relative humidity in the climate-controlled chamber were (mean ± SD) 18.9 ± 2.9 °C and 46.7 ± 11.9%, respectively. The mean O_3_ concentrations for the FA and O_3_ exposures were 14.5 ± 3 ppb and 160.7 ± 5 ppb, respectively (see Supplemental Table 2).

### Ozone-induced Changes in Spirometric Indices

The mean pre and post-exercise spirometric values for FEV_1_, FVC, and FEV_1_/FVC are shown in Supplemental Table 3 and Figure 1. FA exposure did not cause any significant change in FEV_1_ or FVC. By contrast, O_3_ exposure caused a significant decline in FVC (p=0.005) and a non-signifiacnt decline in FEV_1_ (p=0.094); these differences between the FA and O_3_ exposures were statistically significant (Figure 1A). No statistically significant differences were seen 18 h after the two types of exposure, prior to the challenge bronchoscopies. There were also no differences in lung function response to O_3_ between GSTM1-present and GSTM1-null participants.

**Figure 1.**
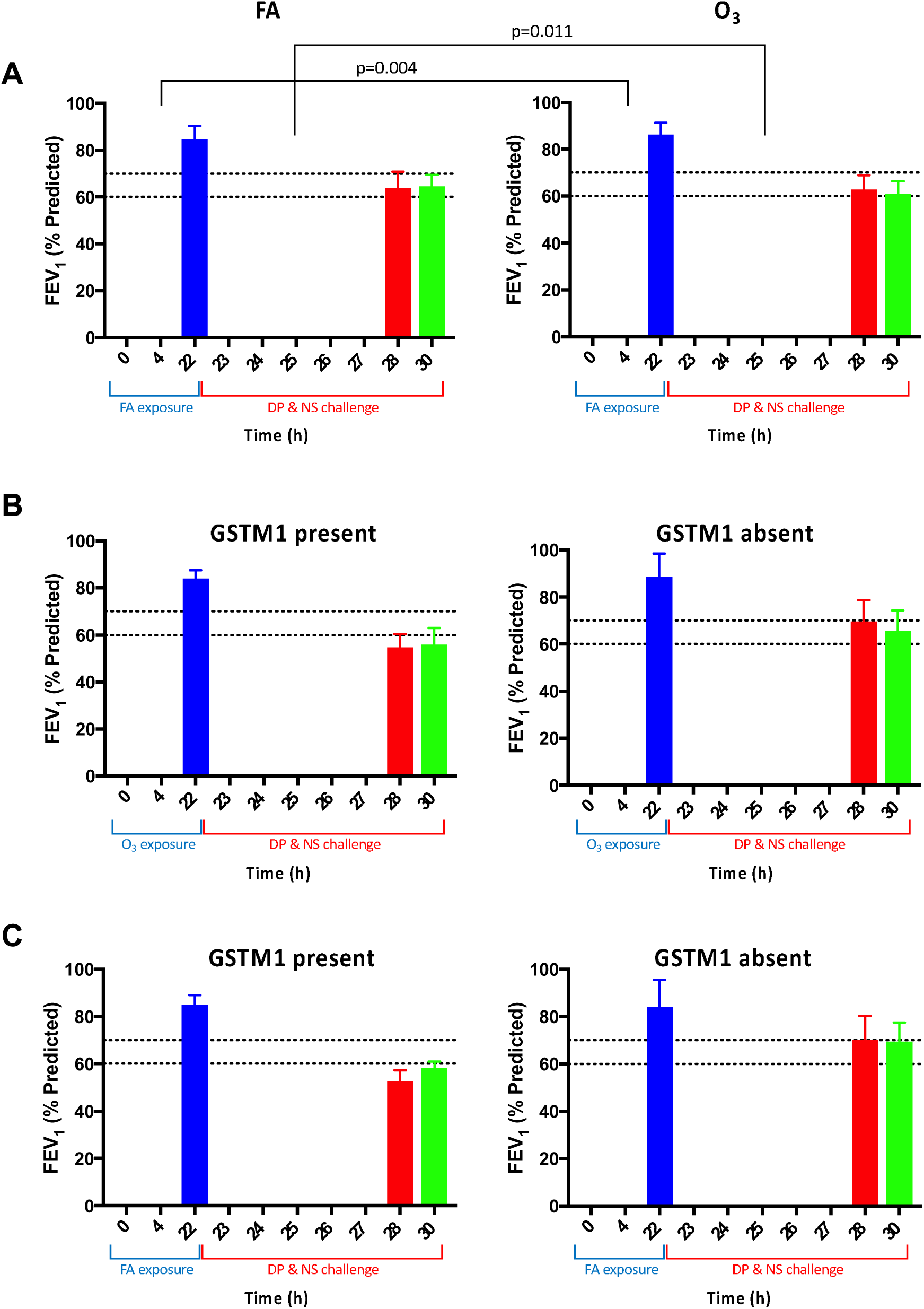
Lung function changes across ozone (O_3_) or filtered air (FA) exposure followed by *Dermatophagoides pteronyssinus* (DP) allergen and normal saline control (NS) challenge via local endobronchial allergen challenge (LEAC). Changes in forced expiratory volume in 1 second (FEV_1_) as a percent predicted of normal values over time (in hours, h) are shown. Blue shade bars show the FEV_1_ across FA or O_3_ exposure. Red shade bars show FEV_1_ after LEAC. Green bar shows FEV_1_ after sampling bronchoscopy with bronchoalveolar lavage (BAL). Row A shows comparison between FA and O_3_ exposure. Rows B and C show comparisons of FEV_1_ response between GSTM1 present and absent participants after O_3_ and FA exposures, respectively.

### Endobronchial Allergen Challenge-induced Changes in Spirometric Indices

The mean post-LEAC hourly spirometric indices are shown in Figure 1. LEAC caused a signficaint decline in FEV_1_ and FVC beginning 1 h post-LEAC. At 3 h post-LEAC, the magnitude of decrease in FEV_1_ was significantly greater after O_3_ by (mean+/- SEM) 10.0 +/- 3.2 percent predicted compared to after FA (p=0.002); the actual difference between FEV_1_ response at 3 h post-LEAC was 6.7 +/- 3.3 % percent predicted lower after O_3_ compared to after FA (p=0.011). At 6 h post-LEAC, the magnitude of decrease in FEV_1_ was significantly greater in participants with GSTM1 present compared to those with GSTM1 absent (mean+/- SEM of 15.7 +/- 5.2 % predicted; p=0.008). However, O_3_ exposure did not cause any significant difference in the FEV_1_ response at 6 h post-LEAC.

### Ozone- and Allergen-induced Changes in BAL Inflammatory Cell Indices

Bronchoalveolar lavage cellular data are shown in Figure 2. Independent of O_3_ exposure, DP challenge compared to saline challenge caused a significant BAL leukocytosis (p=0.02) mainly due to increased eosinophils (p<0.001) and lymphocytes (p<0.005). There was also a non-significant trend towards increased neutrophils (p=0.11), which seemed to be mainly due to the neutrophilic response in partcipants with the GSTM1-present genotype (p=0.09 in wildtype versus p=0.879 in null). BAL macrophage counts did not significantly change. Independent of allergen challenge, O_3_ exposure on its own did not cause any changes in BAL total cells or cell composition.

**Figure 2.**
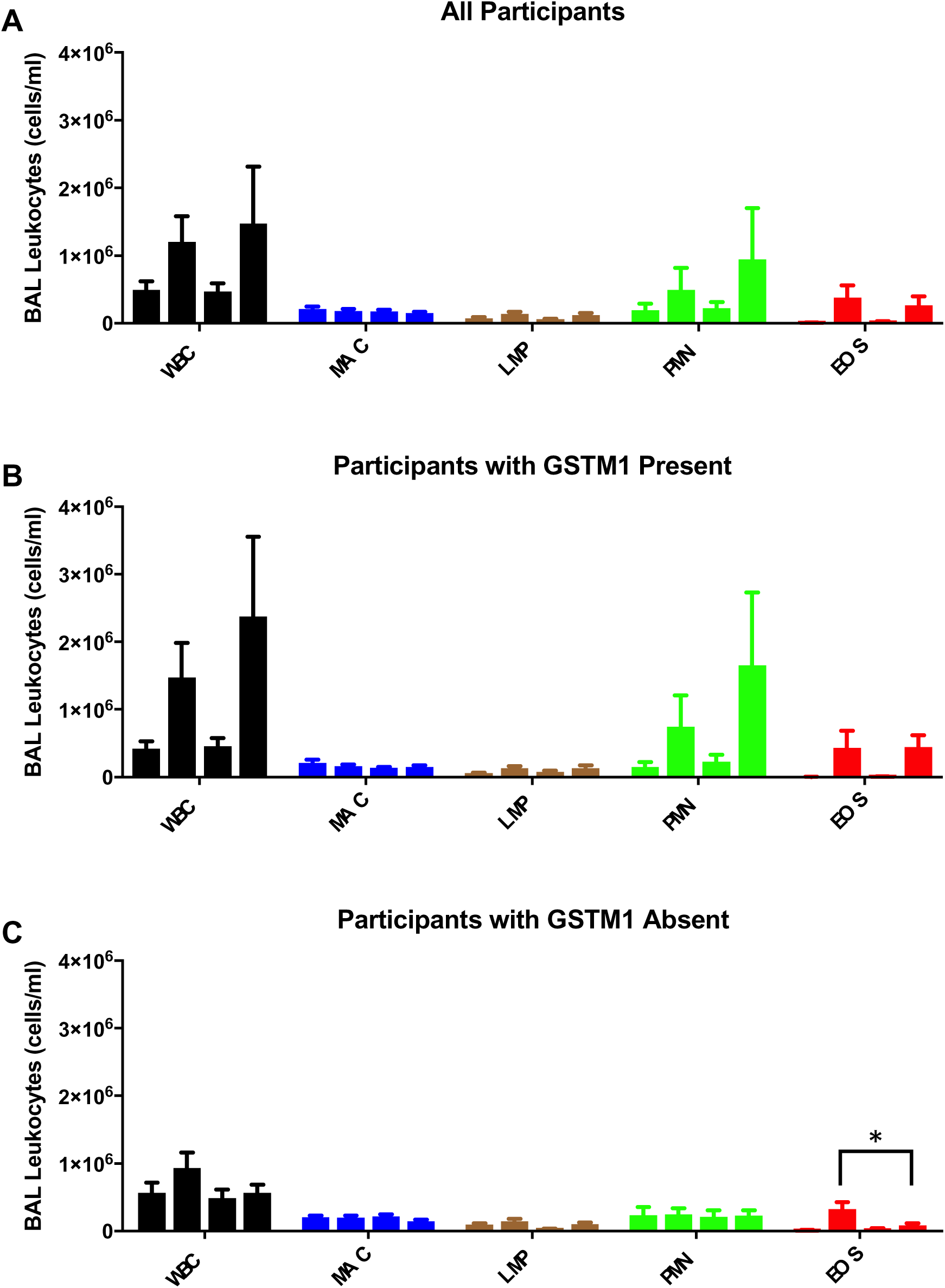
Bar plots (mean and standard error of mean) of cell concentrations in bronchoalveolar lavage (BAL) fluid obtained 6 h after local endobronchial allergen challenge. Panel A, all subjects; panel B, GSTM1 present participants; panel C GSMT1 absent participants. DP=*Dermatophagoides pteronyssinus* antigen; FA=filtered air; NS=normal saline control; O_3_=ozone; GSTM1 null=glutathione-S-transferase mu1 null genotype; GSTM1 WT=glutathione-S-transferase mu1 present genotype; Left to right histograms for each color-coded cell type: FA-NS, O_3_-NS, FA-DP, O_3_-DP. Short bars indicate median values.

Overall, O_3_ exposure combined with DP allergen challenge did not cause any changes in BAL total cells or cell composition. However, in partcipants with the GSTM1-null genotype, O_3_ exposure caused a significant attenuation of the BAL eosinophil response after DP challenge (p=0.041), but not in participants with the GSTM1-present genotype. GSTM1 genotype had no significant effect on the BAL counts of other cell types.

### Ozone- and Allergen-induced Changes in BAL Inflammatory Cytokine Indices

Bronchoalveolar lavage cytokine data are shown in Figure 3. Independent of O_3_ exposure, allergen challenge compared to saline challenge caused a significant increase in BAL concentrations of Th2 cytokines including IL-4, IL-5, IL-10, and IL-13 (P≤0.026 for all comparisons), but no significant change in Th1 cytokines (Il-1b, IL-6, IL-8, TNF-α, or GM-CSF). Independent of allergen challenge, O_3_ exposure did not cause any changes in Th1 or Th2 cytokines.

**Figure 3.**
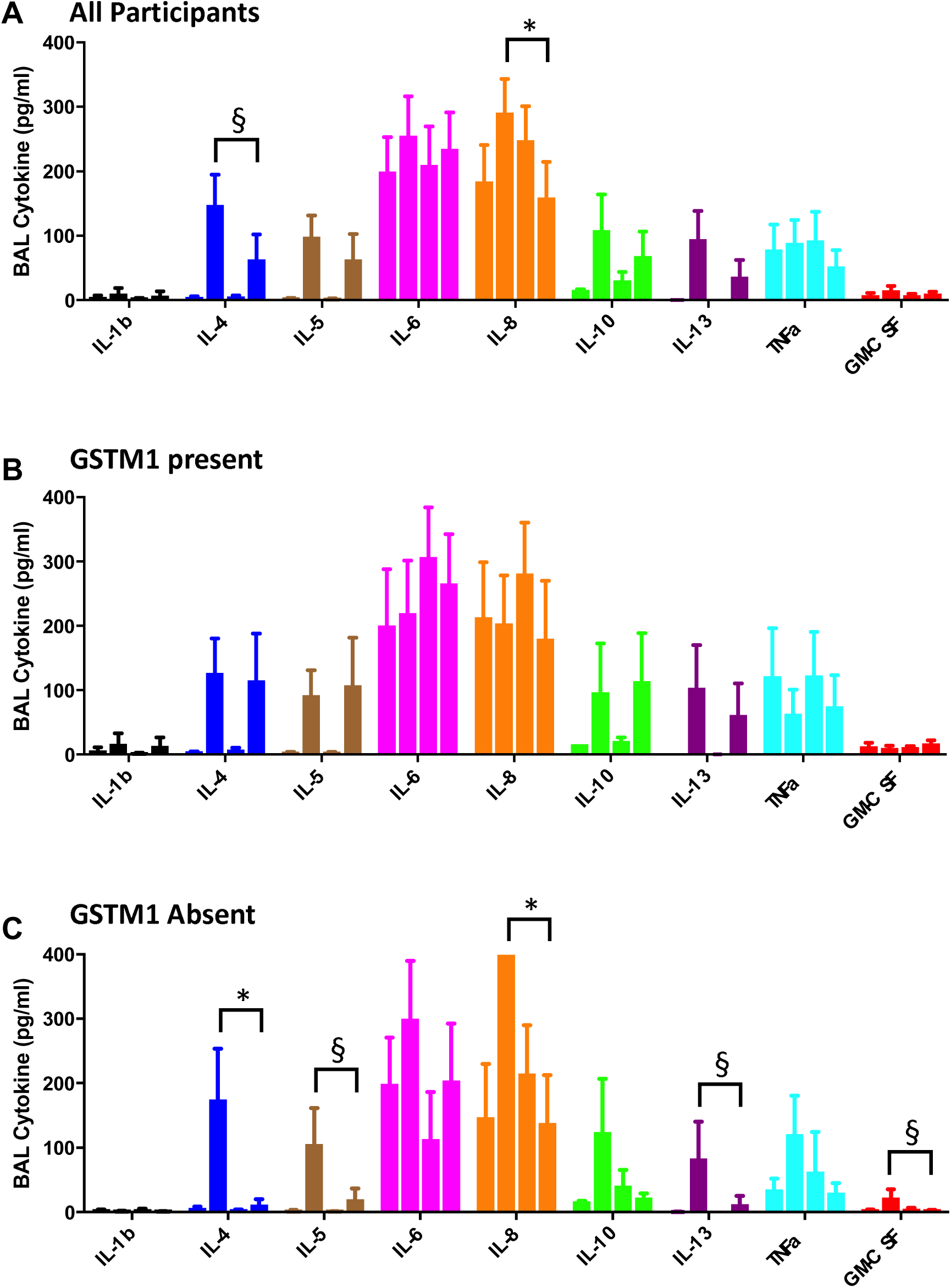
Bar plots (mean and standard error of mean) of cytokine concentrations in bronchoalveolar lavage (BAL) fluid obtained 6 h after local endobronchial allergen challenge (LEAC). Panel A, all participants; panel B, participants with GSTM1 present; panel C participants with GSMT1 absent. The upper limit of detection for IL-5, IL-8, and IL-13 was 400 pg/mL. The lower limits of detection were as follows: IL-4, IL-5, IL-8, and GM-CSF: 0.03, IL-10: 16, and IL-13: 0.13 pg/mL. When values were outside of the detection range, the upper and lower limits of detection were used. BAL= bronchoalveolar lavage; DP=*Dermatophagoides pteronyssinus* antigen; FA=filtered air; NS=normal saline control; O_3_=ozone; GSTM1 absent=glutathione-S-transferase mu1 null genotype; GSTM1 present=glutathione-S-transferase mu1 wild type genotype. Left to right histograms for each color-coded cell type: FA-NS, O_3_-NS, FA-DP, O_3_-DP. Short bars without symbols indicate median values. Bars with symbols indicate differences between groups: §=p<0.1, *=p<0.05.

Overall, O_3_ exposure combined with DP allergen challenge caused a significant decrease in BAL IL-8 concentration (p=0.021) and a non-significant decrease in IL-4 (p=0.110), but no significant changes in other BAL cytokine concentrations. However, in participants with the GSTM1-null genotype, and not in participants with the GSTM1-present genotype, O_3_ exposure caused a significant attenuation of the BAL IL-4 concentration after DP challenge (p=0.014). Other BAL Th2 cytokines also showed a similar but non-significant attenuation trend (IL-5 [p=0.088], IL-10 [p=0.088], IL-13 [p=0.152]). Interestingly, Th1 cytokines also showed a similar attenuation signal with the combination of O_3_ exposure and DP challenge in subjects with the GSTM1-null genotype (significant: IL-8 [p=0.007]; non-significant trend: TNF-α [p=0.136] and GM-CSF [p=0.064]).

## DISCUSSION

In this study, we attempted to address the following two questions: 1) whether O_3_ exposure enhances the specific airway inflammatory responses of asthmatic participants during late-phase reactions to inhaled local endobronchial allergen challenge, and 2) whether asthmatic individuals with the GSTM1-null genotype have greater allergic inflammatory responses than those who have GSTM1 present. Our results suggest that O_3_, at least at the concentration (160 ppb) and exposure duration (4 h) tested, appears to have mixed effects on allergen-induced airway inflammation. While there were no significant changes in BAL total cells or cell composition after O_3_-allergen exposure compared to FA- allergen exposure, BAL concentrations of most cytokines assayed were non-significantly lower after O_3_-allergen exposure; IL-8 was significantly lower. Remarkably, the absence of GSTM1 appears to be associated with decreased magnitude of the inflammatory response to endobronchial allergen challenge after O_3_ exposure with attenuation of allergic cells (eosinophils) and both Th2 (IL-4) and Th1 (IL-8) cytokines. These results must be interpreted with caution, given our small sample size. Despite the small sample size, however, we did find that O_3_ exposure significantly enhanced the lung function response to allergen at 3 h after local endobronchial challenge, consistent with previously published studies that used whole lung inhalation challenge (8, 9).

As expected from previous research in our laboratory and elsewhere, O_3_ exposure did induce a significant but temporary decrease in lung function (2, 4, 25). The mechanism underlying the significant enhancement by O_3_ exposure of the bronchoconstrictor response to allergen at 3 h after local endobronchial challenge is probably enhanced local bronchoconstriction of the allergen-challenged lung segment. We directly observed narrowing of the lumen of the previously challenged segment at the time of the sampling bronchoscopy 6 h after allergen challenge bronchoscopies following both O_3_ and FA exposures. Ozone exposure itself is known to cause some bronchoconstriction even in non-asthmatic participants, possibly due to airway edema and/or neuroreceptor stimulation (26). It is likely that the direct effects of O_3_ on the airways are additive to those of specific allergen challenge. Although previous reports in the literature have suggested that the GSTM1 genotype enhances lung function responses to O_3_ (27–29), we found no evidence for such an effect. In fact, the participants with GSTM1 present had the largest decreases in FEV_1_ and FVC after 4-h exposure to O_3_.

The novel finding of our study, a suggestion that the airway inflammatory cytokine response to specific allergen challenge is decreased after O_3_ exposure, also requires mechanistic explanation. One possibility is that O_3_ exposure leads to activation of innate immunity which may, in turn, dampen Th2 responses to allergen. The results of several studies support such an effect of exposure to an innate immune stimulus, through an IFN-γ-dependent mechanism (30–32) that may involve both a toll-like receptor pathway (32) and lung macrophages (30). However, there is also evidence that O_3_ activation of innate immunity actually enhances Th2 responses (33). Other investigators have found evidence of IL-8 involvement in the late-phase inflammatory response to allergen in sensitized participants (34). Thus, our finding of a decreased IL-8 cytokine response after O_3_ pre-exposure to allergen in GSTM1-null partcipants is intriguing and perhaps consistent with the decreased Th2 cytokine responses to allergen after O_3_ pre-exposure in these participants.

We also found no evidence of an enhanced airway neutrophilic inflammatory response after O_3_-allergen exposure in the GSTM1-null participants. To our surprise, the GSTM1-null participants had lower airway cellular and cytokine responses to O_3_-allergen exposure than GSTM1-present participants. We had hypothesized that GSTM1-null participants would experience greater oxidative stress after O_3_ pre-exposure than GSTM1-present participants and thus would have greater airway cellular and cytokine inflammatory responses to subsequent allergen challenge. Although we actually found a suggestion of a decreased airway inflammatory response to allergen after O_3_ pre-exposure in the GSTM1-null participants, this finding should be considered preliminary until confirmed in another study.

Our study has both strengths and limitations. It is the first controlled human exposure study of an air pollutant to use local endobronchial allergen challenge followed by measurement of biomarkers of airway inflammation in BAL. It is also the first study to assess the impact of the common GSTM1 null genetic variant on airway responses to allergen after O_3_ exposure. Limitations include relative lack of power to study small changes (e.g., the trend toward an increase in BAL neutrophils after O_3_-allergen exposure might have become significant with a larger sample size) and the recruitment of participants with relatively mild allergic asthma for safety reasons, given that the effects of O_3_ inhalation on local endobronchial allergen challenge in specifically sensitized asthmatic participants had not been previously studied. It is possible that patients with more severe asthma are at greater risk for O_3_-induced effects on allergic inflammatory responses. Another potential limitation is the simultaneous use of saline and allergen endobronchial challenge in different lobes. To avoid any potential cross contamination of saline and allergen, we performed the saline challenge in RUL and the allergen challenge in RML, and asked the participants to remain in the recumbent position during the period between the LEAC and sampling bronchoscopies. In addition, during the sampling bronchoscopy, we first performed lavage of the RUL followed by lavage of the RML. Nevertheless, it is possible that local allergen challenge contributes to a systemic signal which could affect lung responses at other sites including the site challenged with saline. However, such cross-reactions would only introduce a bias towards not seeing a difference in responses between saline and allergen challenge.

Our results confirm previous reports that O_3_ pre-exposure enhances the lung function response to allergen in specifically sensitized asthmatic individuals. The novel finding of this study, however, is that O_3_ exposure appears to decrease the cytokine component of the airway inflammatory response to allergen in these individuals. Moreover, the absence of the antioxidant enzyme, GSTM1, does not seem to increase the bronchoconstrictor response and may decrease the airway inflammatory response to allergen following O_3_ exposure. Other recent studies also suggest that GSTM1-deficient individuals do not always have enhanced responses to O_3_ exposure (35–37).

## Data Availability

All data produced in the present study are available upon reasonable request to the authors.

## ONLINE SUPPLEMENT

**Supplemental Table 1:** Dermatophagoides pteronyssinus (DP) allergen skin reactions and concentrations used for local endobronchial allergen challenge and the lung function response.

| Participant ID# | GSTM1 status | Dilution of 10,000 AU/mL allergen that caused $\geq 3 \times 3$ mm skin wheal | Dilution of 10,000 AU/mL allergen used for LEAC | Highest Percent Decline in FEV<sub>1</sub> within 6 h after LEAC | Exposure |
| --- | --- | --- | --- | --- | --- |
| 01 | WT | $1 \times 10^{-3}$ | $1 \times 10^{-4}$ | 21.8% | FA |
|  |  |  |  | 20.9% | O<sub>3</sub> |
| 02 | Null | $1 \times 10^{-1}$ | $1 \times 10^{-2}$ | 25.9% | FA |
|  |  |  |  | 36.0% | O<sub>3</sub> |
| 04 | WT | $1 \times 10^{-4}$ | $1 \times 10^{-5}$ | 23.4% | FA |
|  |  |  |  | 28.8% | O<sub>3</sub> |
| 22 | WT | $1 \times 10^{-3}$ | $1 \times 10^{-4}$ | 45.4% | FA |
|  |  |  |  | 49.6% | O<sub>3</sub> |
| 24 | Null | $1 \times 10^{-5}$ | $1 \times 10^{-6}$ | 33.0% | FA |
|  |  |  |  | 34.3% | O<sub>3</sub> |
| 29 | WT | $1 \times 10^{-3}$ | $1 \times 10^{-4}$ | 16.9% | FA |
|  |  |  |  | 13.1% | O<sub>3</sub> |
| 30 | Null | $1 \times 10^{-3}$ | $1 \times 10^{-4}$ | 12.3% | FA |
|  |  |  |  | 13.8% | O<sub>3</sub> |
| 31 | Null | $1 \times 10^{-3}$ | $1 \times 10^{-4}$ | 13.7% | FA |
|  |  |  |  | 18.5% | O<sub>3</sub> |
| 32 | Null | $1 \times 10^{-2}$ | $1 \times 10^{-3}$ | 28.2% | FA |
|  |  |  |  | 45.7% | O<sub>3</sub> |
| 33 | WT | $1 \times 10^0$ | $1 \times 10^{-1}$ | 57.2% | FA |
|  |  |  |  | 63.3% | O<sub>3</sub> |

Abbreviations: FEV<sub>1</sub>=forced expiratory volume in 1 second; LEAC= local endobronchial allergen challenge; FA= filtered air; O<sub>3</sub>=ozone; GSTM1 null=glutathione-S-transferase mu1 null genotype (enzyme absent); GSTM1 WT=glutathione-S-transferase mu1 wild type genotype (enzyme present).

**Supplemental Table 2:**
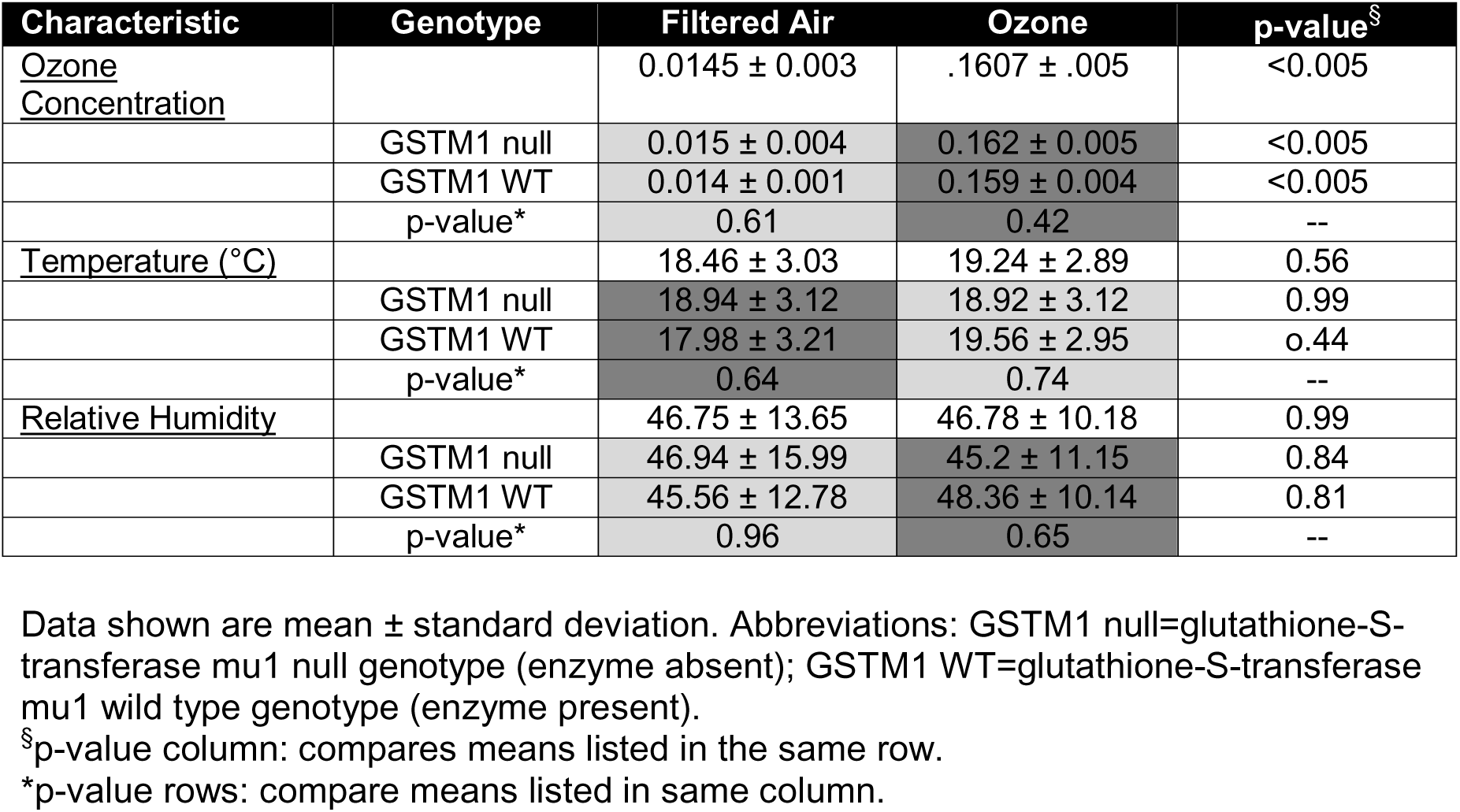
Exposure Characteristics

| Characteristic | Genotype | Filtered Air | Ozone | p-value<sup>§</sup> |
| --- | --- | --- | --- | --- |
| <u>Ozone Concentration</u> |  | 0.0145 ± 0.003 | .1607 ± .005 | <0.005 |
|  | GSTM1 null | 0.015 ± 0.004 | 0.162 ± 0.005 | <0.005 |
|  | GSTM1 WT | 0.014 ± 0.001 | 0.159 ± 0.004 | <0.005 |
|  | p-value* | 0.61 | 0.42 | -- |
| <u>Temperature (°C)</u> |  | 18.46 ± 3.03 | 19.24 ± 2.89 | 0.56 |
|  | GSTM1 null | 18.94 ± 3.12 | 18.92 ± 3.12 | 0.99 |
|  | GSTM1 WT | 17.98 ± 3.21 | 19.56 ± 2.95 | 0.44 |
|  | p-value* | 0.64 | 0.74 | -- |
| <u>Relative Humidity</u> |  | 46.75 ± 13.65 | 46.78 ± 10.18 | 0.99 |
|  | GSTM1 null | 46.94 ± 15.99 | 45.2 ± 11.15 | 0.84 |
|  | GSTM1 WT | 45.56 ± 12.78 | 48.36 ± 10.14 | 0.81 |
|  | p-value* | 0.96 | 0.65 | -- |

Data shown are mean ± standard deviation. Abbreviations: GSTM1 null=glutathione-S-transferase mu1 null genotype (enzyme absent); GSTM1 WT=glutathione-S-transferase mu1 wild type genotype (enzyme present).
<sup>§</sup>p-value column: compares means listed in the same row.
\*p-value rows: compare means listed in same column.

**Supplemental Table 3:** Serial measurement of lung function: comparison of ozone-allergen vs. filtered air-allergen

| Time (h) | Lung Function | Filtered Air | Ozone | p-Value |
| --- | --- | --- | --- | --- |
| 0 (Pre-exposure) | FEV<sub>1</sub> | 3.15 ± .21 | 3.11 ± .18 | 0.63 |
|  | FVC | 4.25 ± .24 | 4.22 ± .21 | 0.71 |
|  | Ratio<br>FEV<sub>1</sub>/FVC | 0.75 ± .03 | 0.74 ± .03 | 0.84 |
| 4 (Post-exposure) | FEV<sub>1</sub> | 3.22 ± .22 | 3.00 ± .21 | **0.03** |
|  | FVC | 4.25 ± .23 | 4.03 ± .23 | **0.03** |
|  | Ratio<br>FEV<sub>1</sub>/FVC | 0.76 ± .03 | 0.75 ± .03 | 0.24 |
| 22 (Pre-LEAC) | FEV<sub>1</sub> | 3.04 ± .21 | 3.12 ± .18 | 0.33 |
|  | FVC | 4.05 ± .24 | 4.18 ± .22 | 0.21 |
|  | Ratio<br>FEV<sub>1</sub>/FVC | 0.75 ± .03 | 0.75 ± .03 | 0.78 |
| 22 (Pre-LEAC) * | FEV<sub>1</sub> | 2.87 ± .19 | 2.98 ± .16 | 0.25 |
|  | FVC | 3.84 ± .18 | 3.99 ± .21 | 0.09 |
|  | Ratio<br>FEV<sub>1</sub>/FVC | 0.75 ± .03 | 0.75 ± .03 | 1.00 |
| 23 (1 h Post-LEAC) * | FEV<sub>1</sub> | 2.55 ± .17 | 2.44 ± .21 | 0.18 |
|  | FVC | 3.43 ± .20 | 3.21 ± .25 | 0.18 |
|  | Ratio<br>FEV<sub>1</sub>/FVC | 0.77 ± .03 | 0.76 ± .03 | 0.65 |
| 24 (2 h Post-LEAC) * | FEV<sub>1</sub> | 2.55 ± .18 | 2.31 ± .17 | 0.08 |
|  | FVC | 3.36 ± .23 | 3.01 ± .26 | 0.20 |
|  | Ratio<br>FEV<sub>1</sub>/FVC | 0.76 ± .03 | 0.77 ± .03 | 0.78 |
| 25 (3 h Post-LEAC) * | FEV<sub>1</sub> | 2.57 ± .18 | 2.30 ± .19 | **0.04** |
|  | FVC | 3.46 ± .25 | 3.02 ± .24 | 0.13 |
|  | Ratio<br>FEV<sub>1</sub>/FVC | 0.75 ± .04 | 0.77 ± .03 | 0.61 |
| 26 (4 h Post-LEAC) * | FEV<sub>1</sub> | 2.37 ± .23 | 2.46 ± .24 | 0.41 |
|  | FVC | 3.33 ± .36 | 3.32 ± .29 | 0.91 |
|  | Ratio<br>FEV<sub>1</sub>/FVC | 0.72 ± .04 | 0.75 ± .04 | 0.47 |
| 27 (5 h Post-LEAC) * | FEV<sub>1</sub> | 2.42 ± .17 | 2.36 ± .26 | 0.67 |
|  | FVC | 3.31 ± .22 | 3.23 ± .26 | 0.60 |
|  | Ratio<br>FEV<sub>1</sub>/FVC | 0.73 ± .03 | 0.72 ± .04 | 0.73 |
| 28 (6 h Post-LEAC) * | FEV<sub>1</sub> | 2.30 ± .21 | 2.29 ± .27 | 0.87 |
| (Pre-Sampling<br>Bronchoscopy) | FVC | 3.11 ± .25 | 3.11 ± .32 | 0.98 |
|  | Ratio<br>FEV<sub>1</sub>/FVC | 0.74 ± 0.4 | 0.74 ± .03 | 0.83 |

Data shown are mean ± standard error of mean (SEM). Abbreviations: LEAC= local endobronchial allergen challenge; FEV<sub>1</sub>=forced expiratory volume in 1 second; FVC=forced vital capacity.
\*spirometry values obtained using EasyOne spirometer; LEAC=local endobronchial allergen challenge.

**Supplemental Table 4:**
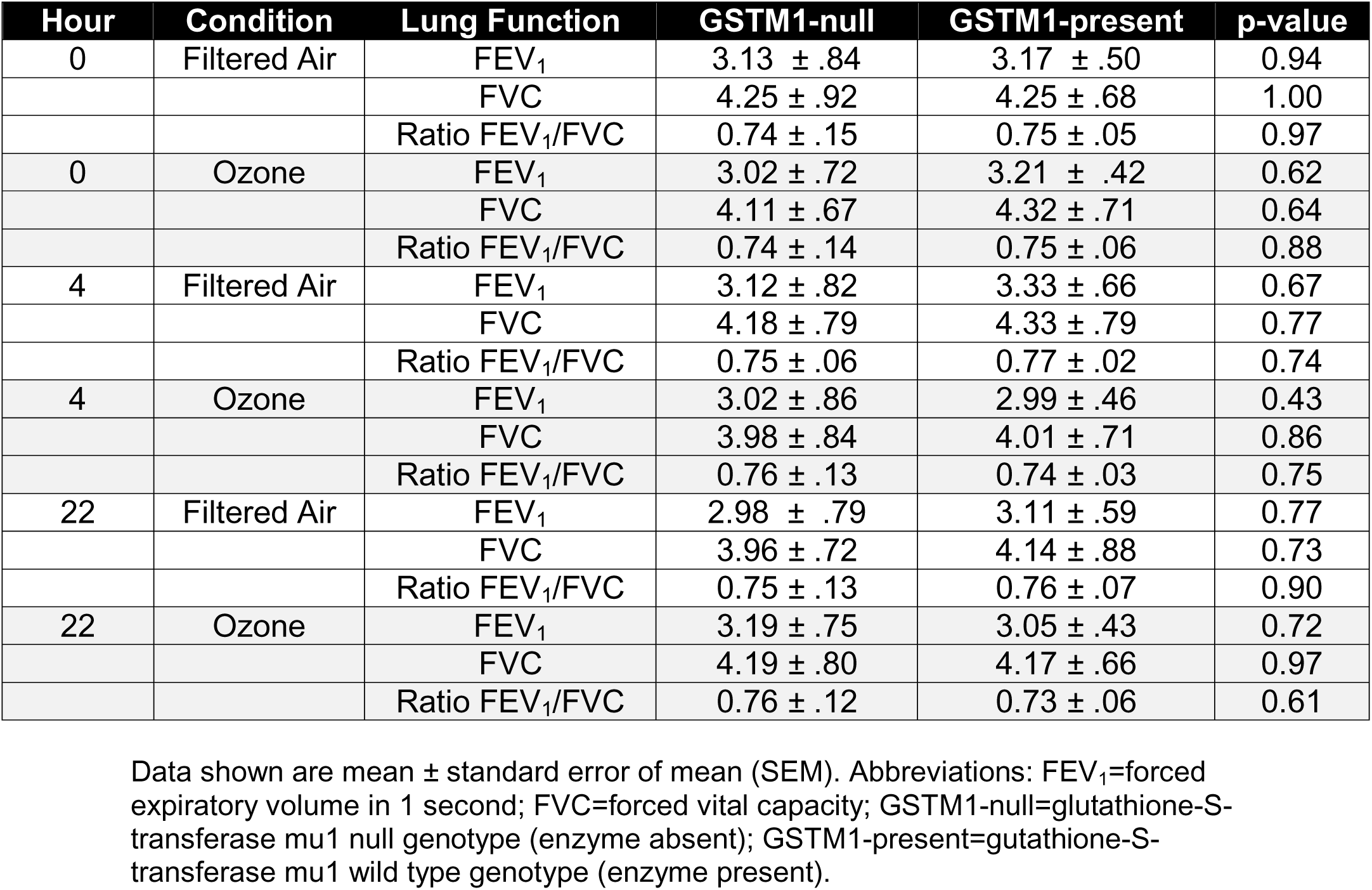
Serial measurement of lung function across ozone and filtered air exposures: comparison of GSTM1-null vs. GSTM1-present genotypes

